# Are college campuses superspreaders? A data-driven modeling study

**DOI:** 10.1101/2020.12.18.20248490

**Authors:** Hannah Lu, Cortney Weintz, Joseph Pace, Dhiraj Indana, Kevin Linka, Ellen Kuhl

## Abstract

The COVID-19 pandemic continues to present enormous challenges for colleges and universities and strategies for save reopening remain a topic of ongoing debate. Many institutions that reopened cautiously in the fall experienced a massive wave of infections and colleges were soon declared as the new hotspots of the pandemic. However, the precise effects of college outbreaks on their immediate neighborhood remain largely unknown. Here we show that the first two weeks of instruction present a high-risk period for campus outbreaks and that these outbreaks tend to spread into the neighboring communities. By integrating a classical mathematical epidemiology model and Bayesian learning, we learned the dynamic reproduction number for 30 colleges from their daily case reports. Of these 30 institutions, 14 displayed a spike of infections within the first two weeks of class, with peak seven-day incidences well above 1,000 per 100,000, an order of magnitude larger than the nation-wide peaks of 70 and 150 during the first and second waves of the pandemic. While most colleges were able to rapidly reduce the number of new infections, many failed to control the spread of the virus beyond their own campus: Within only two weeks, 17 campus outbreaks translated directly into peaks of infection within their home counties. These findings suggests that college campuses are at risk to develop an extreme incidence of COVID-19 and become superspreaders for neighboring communities. We anticipate that tight test-trace-quarantine strategies, flexible transition to online instruction, and–most importantly–compliance with local regulations will be critical to ensure a safe campus reopening after the winter break.

## 1. Introduction

Almost 400,000 cases of COVID-19 have been reported across 1,800 colleges and universities in the United States since the beginning of the pandemic in spring 2020 (New York Times 2020). On March 6, 2020, Stanford University and Touro College became the first institutions nationwide to entirely switch to online instruction to manage the spread of COVID-19 (Kadvany 2020). By the end of the week, more than half of all degree-granting private non-profit and public four-year institutions announced their transition to online learning (Marsicano et al. 2020). Nearly 1,400 colleges and universities delivered their courses online by the end of March and most institutions remained closed throughout the summer (College Crisis Initiative 2020). Many institutions that reopened in the fall have seen massive surges once students returned to campus, some quickly returned to online instruction, others have not yet reopened to date (Chronicle of Higher Education 2020). Understandably, the question whether and how to re-open schools, colleges, and universities remains an ongoing debate (Andersen et al. 2020). Not only student well-being, quality of education, and public health, but also revenue for institutions and their local economies are heavily affected by the decision and timing of campus reopening (Cheng et al. 2020). While early policy makers had hoped to create local bubbles of a COVID-19 free campus environment, we now know that the virus spreads rapidly among students and that local campus outbreaks are often linked to spiking case numbers in neighboring communities (Ivory et al. 2020). With the end of the fall term, a long winter break, and uncertainties around the coming spring, there is an urgent need to thoroughly review the experience from the fall, systematically learn universal trends, and carefully assess the risks of campus reopening.

After the summer break, most colleges had implemented a mandatory 14-day quarantine period after move-in date, followed by an aggressive weekly, or even twice per week, surveillance testing program to minimize the spread of COVID-19 (Chronicle of Higher Education 2020). To increase transparency, many institutions have shared their test results on public COVID-19 dashboards, most of them updated weekly, some even daily (College Crisis Initiative 2020). Despite best efforts, the reported data are sparse, noisy, fluctuating, and often inconsistent. Interpreting the data with a purely machine-learning based approach would likely result in ill-posed problems and overfitting (Alber et al. 2019). To constrain the parameter space, we propose a data-driven modeling approach in which we combine a classical mathematical epidemiology model with Bayesian learning (Linka et al. 2020). Specifically, we use a susceptible-exposed-infectious-recovered compartment model and learn the dynamics of the effective reproduction number for 30 college campuses from the daily case reports using Bayesian inference (Linka et al. 2020a). Figure 1 illustrates the 30 institutions of our analysis and their reported total case numbers since the beginning of the outbreak ranging from 5,806 at the Ohio State University to 141 at Carnegie Mellon University (New York Times 2020). From the learnt reproduction dynamics, we identify trends in campus-wide outbreak dynamics, discuss the effects of online, hybrid, and in person instruction, and make direct comparisons with the case data of each institution’s home county. Our objective is to identify universal features of a campus outbreak, learn patters of infection and reproduction, perform direct comparisons with each institution’s home county, and make informed recommendations about campus reopening after the winter break.

**Figure 1.**
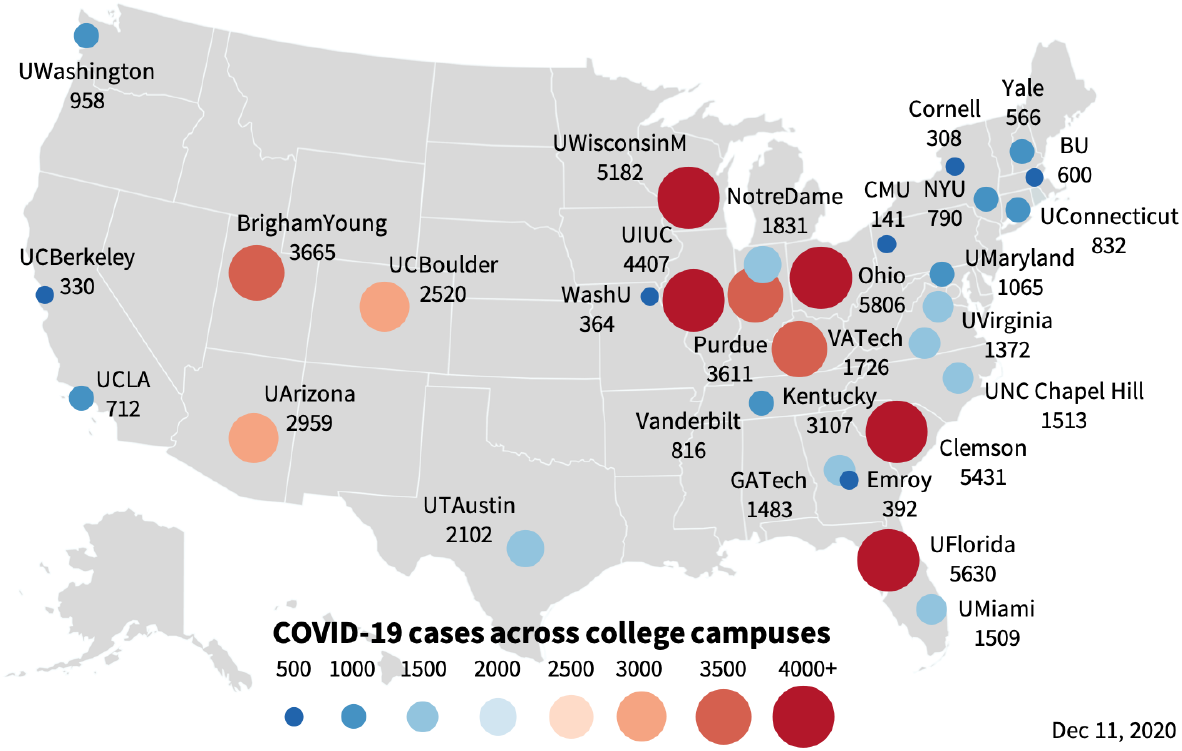
COVID-19 cases across 30 college campuses. Reported cases for ten high case number, public, and private institutions across the United States since the outbreak of the pandemic.

## 2. Methods

### 2.1. COVID-19 college campus data

We drew COVID-19 case reports from 30 publicly available college dashboards across the United States throughout the fall of 2020. We selected 30 institutions for which case numbers are reported on a daily basis and the total cumulative case number exceeds 100; ten institutions with the highest case numbers nationwide (New York Times 2020) and ten public and ten private institutions motivated by national rankings (U.S. News & World Report 2020). For direct comparison, we drew the COVID-19 case reports for the home county of each institution (USA Facts 2020). Table 1 summarizes the reported total enrollment (Chronicle of Higher Education 2020), type of instruction (Chronicle of Higher Education 2020), maximum seven-day per 100,000 incidence, total cases (New York Times 2020), and maximum daily cases for all 30 institutions throughout the fall of 2020.

**Table 1.** COVID-19 outbreak dynamics for 30 college campuses. Total enrollment, type or instruction, maximum seven-day per 100,000 incidence, total cases, maximum daily cases, maximum reproduction number R_max_, initial reproduction number R_0_, recovered fraction *R*, days between the beginning of class and maximum campus incidence, and days between maximum campus and county incidence for ten high case number, public, and private institutions across the United States throughout the fall of 2020; ⋆ no data available to infer R_0_; ⋆⋆ county maximum proceeds campus maximum.

|  | total<br>enroll<br>ment | type<br>of in-<br>struction | max<br>inci-<br>dence | total<br>cases | max<br>daily<br>cases | max<br>repro-<br>duction | initial<br>repro-<br>duction | recov-<br>ered<br>fraction | days to<br>campus<br>peak | days to<br>county<br>peak |
| --- | --- | --- | --- | --- | --- | --- | --- | --- | --- | --- |
| **high case number institutions** |  |  |  |  |  |  |  |  |  |  |
| **U Arizona** | 44097 | online | 2700 | 2959 | 261 | 9.04 | 3.87 | 0.07 | 23 | 6 |
| **Clemson** | 24951 | online | 2685 | 5431 | 185 | 1.83 | 0.27 | 0.22 | 9 | 5 |
| **U Wisconsin Madison** | 43463 | online | 2001 | 5182 | 290 | 2.89 | 1.75 | 0.12 | 8 | 4 |
| **UIUC** | 49702 | hybrid | 1710 | 4407 | 230 | 3.08 | 0.15 | 0.09 | 11 | 3 |
| **Ohio State** | 61170 | hybrid | 1619 | 5806 | 284 | 2.84 | 1.39 | 0.09 | 11 | 21 |
| **Brigham Young** | 34499 | in person | 1522 | 3665 | 108 | 1.77 | * | 0.11 | 23 | 4 |
| **U Florida** | 52218 | online | 1494 | 5630 | 174 | 1.88 | 0.22 | 0.11 | 11 | 17 |
| **UC Boulder** | 36681 | hybrid | 1292 | 2520 | 130 | 4.16 | 1.02 | 0.07 | 28 | 4 |
| **U Kentucky** | 29182 | online | 1276 | 3107 | 102 | 2.36 | * | 0.11 | 29 | 2 |
| **Purdue** | 44474 | in person | 1127 | 3611 | 116 | 3.82 | 2.19 | 0.08 | 92 | 4 |
| **public institutions** |  |  |  |  |  |  |  |  |  |  |
| **UNC Chapel Hill** | 30011 | online | 1817 | 1513 | 91 | 2.77 | * | 0.05 | 14 | 1 |
| **Georgia Tech** | 32723 | hybrid | 1104 | 1483 | 109 | 5.76 | 2.92 | 0.05 | 11 | 11 |
| **Virginia Tech** | 34683 | in person | 1102 | 1726 | 84 | 7.60 | 6.56 | 0.05 | 11 | ** |
| **U Virginia** | 24639 | online | 870 | 1372 | 59 | 2.38 | 1.32 | 0.06 | 28 | 10 |
| **U Connecticut** | 27412 | online | 797 | 832 | 56 | 5.33 | 0.87 | 0.03 | 80 | 7 |
| **U Maryland** | 41200 | online | 720 | 1065 | 87 | 2.32 | 1.77 | 0.03 | 9 | ** |
| **UT Austin** | 51832 | hybrid | 704 | 2102 | 117 | 2.81 | 0.11 | 0.04 | 11 | 7 |
| **UCLA** | 44537 | online | 504 | 712 | 59 | 3.41 | 1.04 | 0.02 | 60 | ** |
| **U Washington** | 47400 | online | 409 | 958 | 76 | 10.75 | 1.96 | 0.02 | 4 | ** |
| **UC Berkeley** | 42501 | online | 146 | 330 | 16 | 4.01 | 1.23 | 0.01 | 105 | 16 |
| **private institutions** |  |  |  |  |  |  |  |  |  |  |
| **Notre Dame** | 12607 | in person | 3083 | 1831 | 104 | 3.29 | 0.31 | 0.15 | 13 | ** |
| **U Miami** | 17331 | in person | 1746 | 1509 | 75 | 3.10 | 0.19 | 0.09 | 94 | 3 |
| **Yale** | 13433 | online | 1105 | 566 | 22 | 2.95 | 0.90 | 0.04 | 78 | 3 |
| **Vanderbilt** | 12824 | in person | 990 | 816 | 39 | 2.91 | 0.82 | 0.06 | 80 | ** |
| **Boston University** | 34657 | hybrid | 393 | 600 | 31 | 3.16 | 1.29 | 0.02 | 94 | 8 |
| **Washington St Louis** | 15852 | online | 386 | 364 | 14 | 3.45 | * | 0.02 | 70 | 24 |
| **Emory** | 14458 | online | 337 | 392 | 16 | 2.00 | 0.53 | 0.03 | 95 | 12 |
| **Cornell** | 23600 | hybrid | 262 | 308 | 14 | 3.62 | 2.29 | 0.01 | 5 | ** |
| **CMU** | 14029 | online | 240 | 141 | 10 | 4.36 | 0.25 | 0.01 | 97 | ** |
| **NYU** | 51847 | hybrid | 214 | 790 | 40 | 5.38 | 0.62 | 0.02 | 82 | ** |

### 2.2. Epidemiology model

We modeled the epidemiology of the COVID-19 outbreak using an SEIR model with four compartments, the susceptible, exposed, infectious, and recovered populations (Hethcote 2000). Their evolution follows a set of ordinary differential equations,

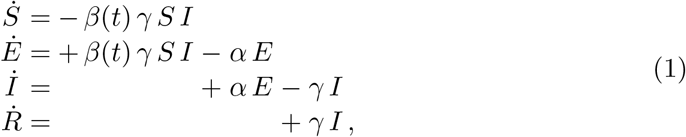

where *α, β*, and *γ* are the latent, contact, and infectious rates. We assumed that the latent and infectious rates, *α* = 2.5/days and *γ* = 6.5/days, are disease-specific and constant (He et al. 2020), and that the contact rate *β*(*t*) is behavior-specific and dynamic (Linka et al. 2020a). We represented the contact rate,

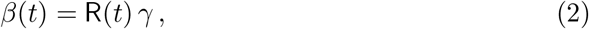

implicitly as the product of the time-varying effective reproduction number R(*t*) and the infectious rate *γ*. We made a Gaussian random walk ansatz for the dynamic reproduction number and assumed a time-varying Gaussian distribution (Peirlinck et al. 2020a),

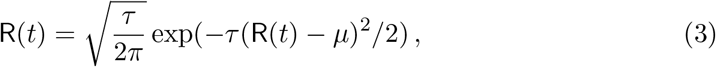

parameterized in terms of the drift *µ* and the daily stepwidth *τ* = *τ*^*∗*^*/*[1.0 − *s*], where *τ*^*∗*^ is the step width precision and *s* is the associated smoothing parameter. We solved the model for the susceptible, exposed, infectious, and recovered populations *S, E, I*, and *R*, and, at the same time, learned the dynamics of the reproduction number R(*t*) from the reported case data of all 30 institutions. From these reproduction dynamics, we calculated the maximum reproduction number R_max_ within the reporting interval and the initial reproduction number R_0_ at the first day of class.

### 2.3. Bayesian inference

Our model is parameterized in terms of a set of three parameters for the SEIR model, θ_SEIR_ = {*E*_0_, *I*_0_, *σ*_I_}, the initial exposed and infectious populations *E*_0_ and *I*_0_ and the likelihood width *σ*_I_, and a set of three parameters for the Gaussian random walk model, θ_Rt_ = { *µ, τ*^*∗*^, *s*}, the drift *µ*, the step width precision *τ*^*∗*^, and the smoothing parameter *s*. We estimate the model parameters θ = θ_SEIR_ ∪ θ_Rt_ using Bayesian inference with Markov Chain Monte Carlo sampling. We adopt a Student’s t-distribution for the likelihood between the new daily reported cases, Δ*Î*(*t*) = *Î*(*t*_n+1_) − *Î*(*t*_n_), and the model predictions, Δ*I*(*t*, θ) = *I*(*t*_n+1_, θ) − *I*(*t*_n_, θ), with a new-case-number-dependent width (Dehning et al. 2020),

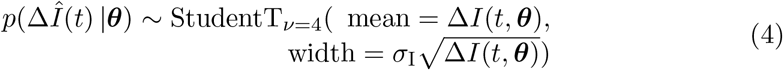

Here, *σ*_I_ represents the width of the likelihood *p*(Δ*Î*(*t*) | θ) between the new daily reported cases Δ*Î*(*t*) and the modeled cases Δ*I*(*t*, θ). We apply Bayes’ rule

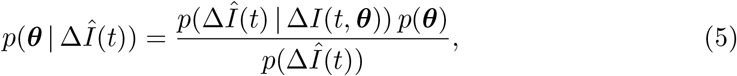

to obtain the posterior distribution of the parameters using the prior distributions from Table 2, and the reported cases themselves, which we infer approximately by employing the NO-U-Turn sampler NUTS (Hoffman & Gelman 2014) implementation of the Python package PyMC3 (Salvatier et al. 2016).

**Table 2.** Bayesian inference. Prior distributions to obtain the posterior distributions of the parameters using Bayes’ rule.

| Parameter | Distribution |
| --- | --- |
| $E_0$ | LogNormal($\log(\Delta I(t = A)), 1.5$) |
| $I_0$ | LogNormal($\log(\Delta I(t = 0)), 1.5$) |
| $\sigma$ | HalfCauchy($\beta = 1$) |
| $R(t)$ | GRW($\mu, \tau_1/(1.0 - s)$) |
| $\mu$ | Normal(0, 2) |
| $\tau^*$ | Exponential(1/2) |
| $s$ | Uniform(0.5, 1) |

## 3. Results

Table 1 summarizes the outbreak dynamics for our ten high case number, public, and private institutions throughout the fall of 2020. The total enrollment, type of instruction, and total cases summarize the reported data; the maximum incidence, maximum daily cases, maximum and initial reproduction numbers, the days between the beginning of class and maximum campus incidence, and days between maximum campus and county incidence summarize the results of our analysis. The type of instruction was primarily online at 16, hybrid at 8, and primarily in person at 6 institutions. The total cases number illustrated in Figure 1 exceeds 5,000 at four institutions, the Ohio State University with 5,806, the University of Florida with 5,630, Clemson University with 5,431, and the University of Wisconsin-Madison with 5,182. The maximum daily case number exceeded 200 at four institutions, the University of Wisconsin-Madison with 290, the Ohio State University with 284, the University of Arizona with 261, and the University of Illinois Urbana Champaign with 230.

Figure 2 illustrates the maximum seven-day per 100,000 incidence, a characteristic metric of the magnitude of the outbreak. The University of Notre Dame faced the highest maximum incidence with 3,083, followed by the University of Arizona with 2,700, and Clemson University with 2,685. According to Table 1, all high case number institutions experienced maxima larger than 1,000, meaning that at least 1% of the student population had tested positive during the peak of the campus outbreak. By the end of the term, the recovered fraction was highest at Clemson University with 22%, followed by the University of Notre Dame with 15%, and the University of Wisconsin-Madison with 12%.

**Figure 2.**
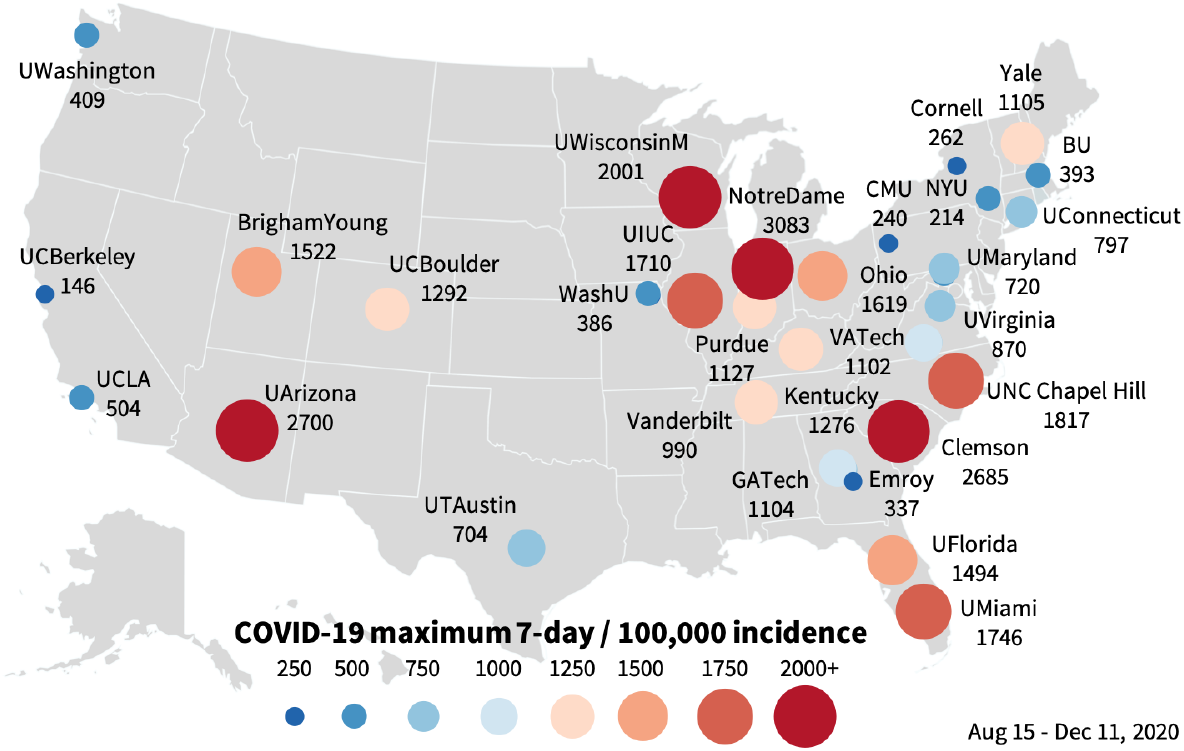
COVID-19 maximum incidence across 30 college campuses. Maximum seven-day per 100,000 incidence of COVID-19 for ten high case number, public, and private institutions across the United States throughout the fall of 2020.

Figure 3 summarizes the COVID-19 outbreak dynamics for the 30 college campuses and their home counties. The circles mark the reported daily campus cases for each institution. The dashed vertical lines mark the first day of class. The red and blue curves highlight the seven-day averages of the daily new campus and county cases, the red and blue dots indicate the timing and peak of their seven-day incidence within the first half of the fall term. Table 1 summarizes the time delays between the beginning of class, indicated through the dashed vertical line, and the maximum seven-day campus incidence, indicated through the red dot. The delay lies within a two-week window for 13 institutions, between three to four weeks at five institutions, and exceeds two months for 12 institutions. The thirteen institutions, five of the high-case group, six public, and two private, displayed a clear maximum of new cases within the first two weeks of the fall term, followed by a rapid drop, and a gradual steady increase towards a second peak at the end of the term. The twelve institutions, one of the high case group, three public, and eight private, displayed no initial peak, but only a gradual increase towards a maximum at the end of the term. Table 1 also quantifies the time delays between the maximum campus and county incidences, indicated through the red and blue dots. The delay lies within a two-week window for 17 institutions, between two to four weeks at four institutions, and for 9 institutions. Strikingly, during the first half of the term, the outbreak dynamics of the home county closely followed the dynamics of most campuses; during the second half, the case numbers of the counties steadily increased, independent of the associated campus dynamics. This similar but shifted trend is particularly visible for the University of Arizona, the University of Wisconsin-Madison, the University of Illinois Urbana Champaign, the Ohio State University, Brigham Young, the University of Colorado Boulder, the University of North Carolina Chapel Hill, the University of Maryland, the University of Texas Austin, the University of Notre Dame, Cornell University, and their home counties.

**Figure 3.**
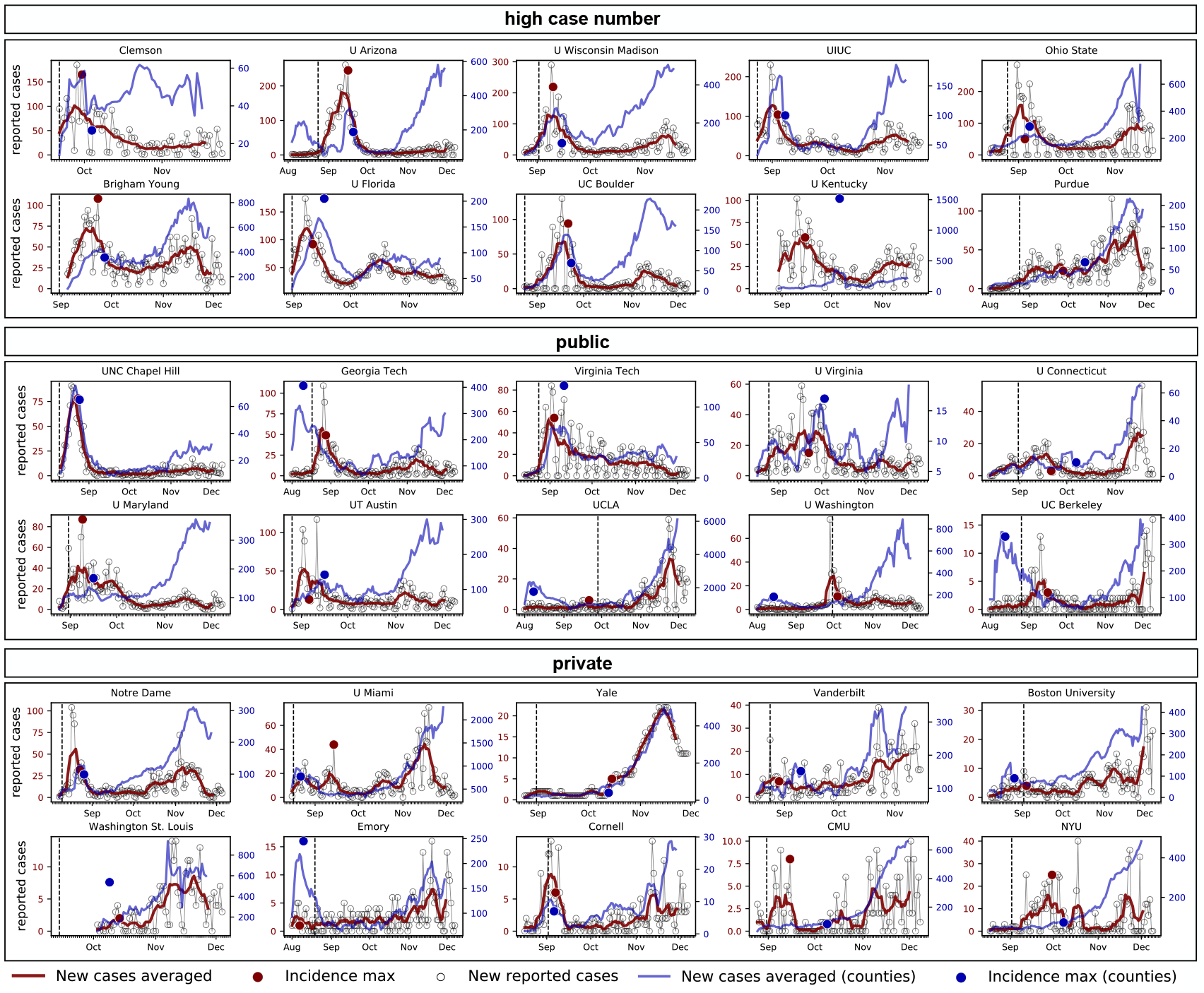
COVID-19 outbreak dynamics for 30 college campuses and their counties. Reported cases and seven-day averages for ten high case number, public, and private institutions across the United States throughout the fall of 2020. Circles mark the reported daily campus cases; dashed vertical lines mark the first day of class; red and blue curves highlight the seven-day averages of daily new campus and county cases; red and blue dots indicate the timing and peak of their seven-day incidence within the first half of the fall.

Figure 4 summarizes the dynamics of the reproduction number R(*t*) inferred from the reported case data using a Gaussian random walk approach. The narrow confidence intervals on both the dynamic reproduction number and the new infectious population suggest that the Gaussian random walk approach is well suited to capture the underlying disease dynamics. It nicely captures the frequent fluctuations associated with rapid policy changes to manage campus-wide outbreaks. Table 1 summarizes the maximum reproduction number R_max_ and the initial reproduction number R_0_ at the beginning of class associated with the 30 red curves. The trends of the reproduction dynamics are similar to the trends in Figure 3 with either a maximum within the first two weeks of the fall term and a subsequent gradual increase, or with a single maximum towards the second half of the term. Most maximum reproduction numbers are on the order of two to three indicative of rapid outbreaks. The initial reproduction numbers R_0_ are consistently lower than the maxima R_max_, most are less than one.

**Figure 4.**
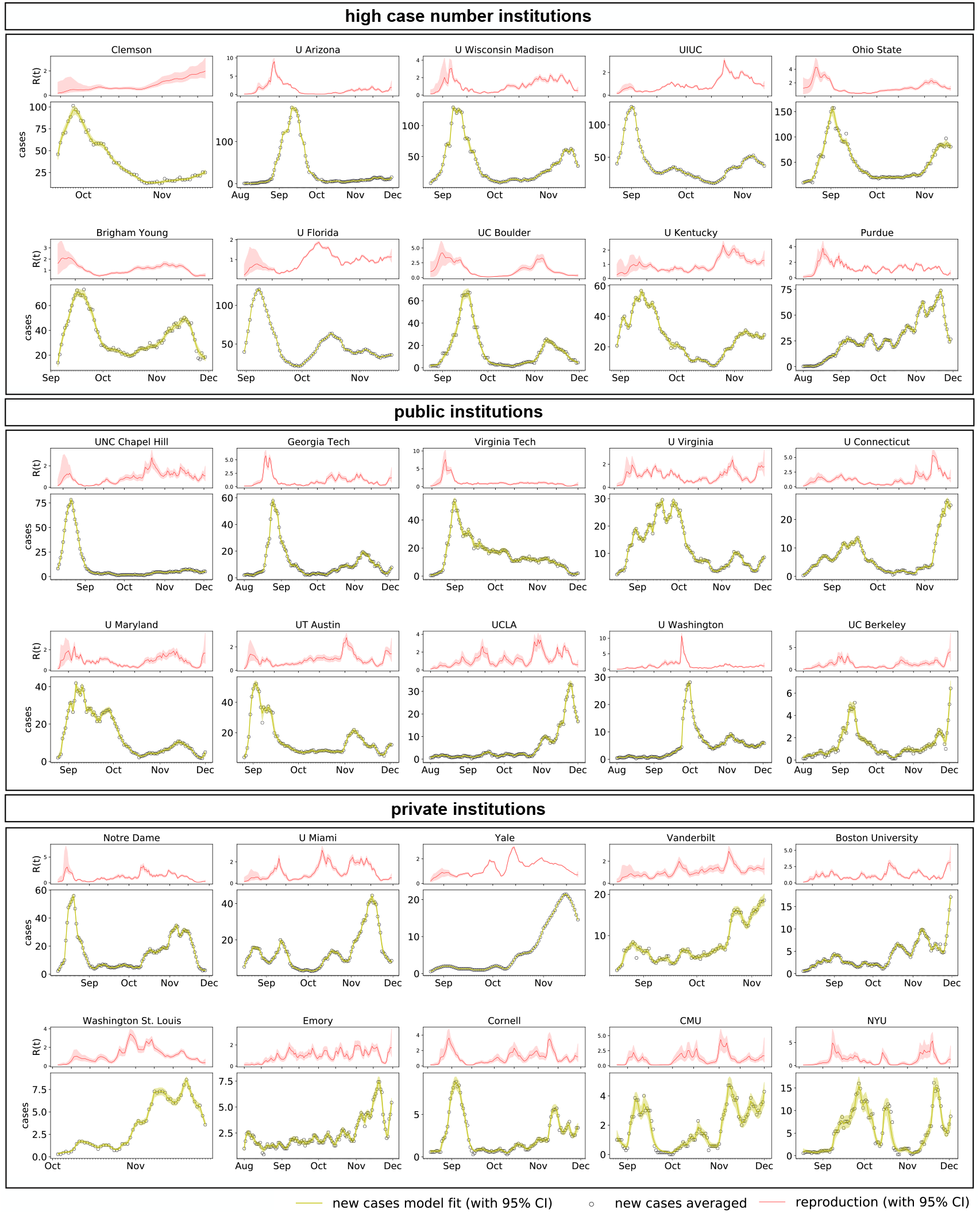
COVID-19 dynamic reproduction number of for 30 college campuses. Dynamic reproduction number, reported cases, and new infectious population for ten high case number, public, and private institutions across the United States throughout the fall of 2020. Circles mark the reported daily campus cases; red and yellow curves highlight the learnt Gaussian random walk based dynamic reproduction number and new infectious population, both with confidence intervals.

## 4. Discussion

The COVID-19 pandemic continues to present enormous challenges for colleges and universities and strategies for safe campus reopening remain a topic of major concern (Cheng et al. 2020). Shortly after the beginning of the fall term, many colleges that had invited their students back to campus experienced a massive spike of new infections (Andersen et al. 2020). This raised the question under which conditions college campuses can reopen safely and to which extent they display superspreading characteristics (Hubler & Hartocollis 2020). Superspreading events–events during which certain individuals infect a disproportionally large number of others–are of particular concern in epidemiology since they are associated with a much faster increase in new cases than spread in a homogeneous population (Lloyd et al. 2005). Identifying superspreading requires sophisticated contact tracing and becomes increasingly difficult when mitigation strategies overwrite the natural outbreak dynamics (Adam et al. 2020). Here, rather than attempting to perform a rigorous statistical analysis of superspreading, we explored reported campus case data and compared the characteristic features of campus-wide outbreaks with their neighboring communities. This allows us to answer the question whether there is a correlation between campus reopening and potential outbreaks, whether college campuses have a higher incidence of COVID-19 than their neighboring communities, and whether college campuses are at risk to become superspreaders.

### The first two weeks of instruction present a high-risk period for campus outbreaks

Out of the 30 colleges and universities of our study, 14 displayed a spike in cases within the first two weeks of class, six with primarily online instruction, six hybrid, and two primarily in person, see Table 1 and Figure 3. For 18 institutions, eight within primarily online instruction, six hybrid, and four primarily in person, the maximum incidence occurred within August 19 and October 10, the interval between the second and third waves of COVID-19 infections, during which the nationwide number of new cases had dropped below 50,000 per day (Johns Hopkins University 2020). This suggests that these initial college outbreaks are unrelated to the national outbreak dynamics. Instead, they are independent local events driven by campus reopening and inviting students back to campus (Marsicano et al. 2020). Our results are a quantitative confirmation of the common fear in early fall that colleges could become the new hot spots of COVID-19 transmission (Hubler & Hartocollis 2020). However, as our results in Figure 4 suggest, most college campuses have successfully responded to these challenges and reduced their reproduction numbers rapidly well below one within a two or three weeks, for example, by temporarily transitioning to online instruction (Robles 2020).

### College campuses experience an extreme incidence of COVID-19

The seven-day per 100,000 incidence is a powerful metric to characterize the outbreak dynamics of COVID-19. It smoothes fluctuations in weekly reporting and scales case numbers to a fixed, easy-to-compare population. Policy decision makers often consider an incidence of 50 as a threshold for high risk counties, states, or countries (Robert Koch Institute 2020). All 30 institutions of our study exceeded this value, three even by two orders of magnitude: The University of Notre Dame experienced a maximum incidence of 3,083, followed by the University of Arizona with 2,700, and Clemson University with 2,685, see Table 1. All colleges and universities, except the University of California, Berkeley, exceeded the peak incidences of the first and second waves in the United States with 70 on April 10, 2020 and 150 on July 25, 2020 (Johns Hopkins University 2020). Even the current third wave incidence of 466 on December 14, 2020 is lower than the peak incidences of 22 of our 30 institutions, see Figure 2. The extreme incidence across college campuses is worrisome and calls for tight and aggressive mitigation strategies (Cheng et al. 2020). As a result, by the end of the fall term, six institutions had a recovered population of more than 10%, ranging from Clemson University with 22%, to the University of Notre Dame with 15%, the University of Wisconsin Madison with 12%, and Brigham Young University, the University of Florida, and the University of Kentucky, all with 11%, see Table 1. This is more than twice of the national average of 5.3%, with 17.3 million reported cases at a population of 328.2 million (Johns Hopkins University 2020). However, with around 90 reported deaths, mainly college employees and not students (New York Times 2020), the campus-related death rate of 0.02% remains well below the average death rate of COVID-19.

### College campus are at high risk to become superspreaders

The University of Notre Dame experienced the highest peak incidence of our study. Notre Dame had tested all 12,607 students before the beginning of class and only nine had tested positive by August 10 when the fall semester started with in person instruction. At this time, the effective reproduction number of R_0_ = 3.29 was well above three. On August 18, after 147 students had tested positive, Notre Dame announced to move all classes online (Robles 2020). Its maximum incidence of 3,083 followed on August 23, less than two weeks after classes had started. At the same time, the state of Indiana experienced minimum infection rates with less than 1,000 daily new cases between the second and third wave (Johns Hopkins University 2020). This suggests that the Notre Dame campus outbreak was a truly local event, unrelated to the dynamics of the home state of Indiana. By the end of the fall semester, 1,831 of the 12,607 enrolled students had become infected, a fraction of 14.5%. At the same time, the cumulative infection rate in St. Joseph County with 21,125 reported cases in a population of 271,826 was 7.8%. This is well above the average of the state of Indiana with 6.5% (USA Facts 2020) and above the national average of 5.3% (Johns Hopkins University 2020), indicating that the initial outbreak at the University of Notre Dame had superspreading-like effects on its home county. This trend is visible in 17 of our 30 institutions, for which the maximum campus incidence triggered a maximum county incidence within a window of two weeks. Our results in Figure 3 suggest that, unlike St. Joseph County, the University of Notre Dame managed to rapidly decrease the spread, contain the virus, and maintain a low incidence throughout the following months. Other institutions, for example the University of North Carolina at Chapel Hill, implemented similar mitigation strategies with equal success. At Chapel Hill, classes started on August 10, primarily in person. The university reported 177 positive cases within the first week of class and moved all undergraduate classes online on August 17 (Robles 2020), the day of minimum new infections statewide (Johns Hopkins University 2020). Exactly two weeks after the beginning of class, the University of North Carolina at Chapel Hill experienced its maximum incidence of 1,817, triggered by an initial reproduction number of R_0_ = 2.77. By the end of the fall semester, 1,513 of the 30,011 enrolled students, had become infected, a fraction of 5.0%. These numbers confirm that the transition to online instruction is a successful strategy for outbreak control. At both institutions, online education resulted in a rapid drop of new infections, with new case numbers steadily below ten by September 3 at Notre Dame and by September 2 at Chapel Hill.

### Limitations

Although we can robustly identify several universal trends, our approach has a few limitations: First, since most colleges report case data on a weekly rather than on a daily basis, the selection of our 30 colleges is likely biased towards institutions that value regular testing and transparent reporting. Second, since the SEIR model is known to perform poorly for low case numbers, we limited our analysis to institutions with more than 100 cases, which may induce an additional implicit bias. Third, since the true on-campus population was often unreported, we approximated the population by the total enrollment, which is likely an overestimate that results in an underestimate of the maximum incidence and recovered fraction. Fourth, instead of accounting for move-in effects through initial conditions, our method collectively represents outbreak dynamics in terms of the time-varying effective reproduction number R(*t*), which could artificially inflate the reproduction in early fall. Last, although we were able to identify universal trends from 30 individual campuses with campus-specific enrollment, living situation, and type of education, its is virtually impossible to exclude neighborhood outbreak dynamics, county- and state-wide mitigation strategies, national holidays and seasonality. This implies that our observed trends are not necessarily a one-to-one predictor for the COVID-19 dynamics upon campus reopening after the winter break.

## 5. Conclusion

Reopening college campuses during the COVID-19 pandemic remains a topic of on-going debate. To identify general trends and make informed recommendations for the spring term, we have analyzed the outbreak dynamics of COVID-19 across 30 campuses and their home counties. All reported campuses pursued regular surveillance testing, weekly or even twice per week, combined with aggressive test-trace-isolate strategies. Tight oversight proved critical, especially during the first two weeks of class, when almost half of the institutions experienced a massive spike in new cases. Strikingly, these local campus outbreaks rapidly spread across the entire county and triggered a peak in new infections in neighboring communities. Interestingly, the majority of colleges and universities were able to rapidly manage their outbreaks and suppress campus-wide infections, while the neighboring communities were less successful in controlling the spread of the virus. As a result, for most institutions, the outbreak dynamics remained manageable throughout the entire fall of 2020 with narrow spikes of less than 300 cases per day. During the second half of the term, case numbers increased gradually towards a second peak, which was universally less pronounced on campuses than across the entire county. Taken together, our study suggests that it is possible for colleges and universities to reopen safely while controlling the spread of COVID-19. Successful reopening relies on limiting the introduction of the virus during the initial weeks of the term, regular testing and rapid tracing, and the collective understanding of the importance of quarantine and isolation. With these strategies in place, our findings suggest that college campuses present a risk to initiate superspreading events, but, at the same time, should be applauded for their rapid responses to successfully manage local outbreaks.

## Data Availability

All data are drawn from public databases. References to these databases are cited in the text and listed in the bibliography.

## Acknowledgments

Hannah Lu, Cortney Weintz, Joseph Pace, Dhiraj Indana contributed equally to this manuscript. This work was a class project of the course ME233 Data-driven modeling of COVID-19, and we acknowledge the support by Amelie Schafer, Oguz Tikenogullari, and Mathias Peirlinck. It was supported by a DAAD Fellowship to Kevin Linka, by a Stanford Bio-X IIP seed grant to Ellen Kuhl, and by the Stanford School of Engineering COVID-19 Research and Assistance Fund.

